# SARS-CoV-2 induces inflammasome-dependent pyroptosis and downmodulation of HLA-DR in human monocytes

**DOI:** 10.1101/2020.08.25.20182055

**Authors:** André C. Ferreira, Vinicius Cardoso Soares, Isaclaudia G. de Azevedo-Quintanilha, Suelen da Silva Gomes Dias, Natalia Fintelman-Rodrigues, Carolina Q. Sacramento, Mayara Mattos, Caroline S. de Freitas, Jairo R. Temerozo, Lívia Teixeira, Eugenio Damaceno Hottz, Ester A Barreto, Camila R. R. Pão, Lohanna Palhinha, Milene Miranda, Dumith Chequer Bou-Habib, Fernando A. Bozza, Patrícia T. Bozza, Thiago Moreno L. Souza

## Abstract

Infection by the severe acute respiratory syndrome coronavirus 2 (SARS-CoV-2) has been associated with leukopenia and uncontrolled inflammatory response in critically ill patients. A better comprehension of SARS-CoV-2-induced monocyte death is essential for the identification of therapies capable to control the hyper-inflammation and reduce viral replication in patients with COVID-19. Here, we show that SARS-CoV-2 induces inflammasome activation and cell death by pyroptosis in human monocytes, experimentally infected and from patients under intensive care. Pyroptosis was dependent on caspase-1 engagement, prior to IL-1ß production and inflammatory cell death. Monocytes exposed to SARS-CoV-2 downregulate HLA-DR, suggesting a potential limitation to orchestrate the immune response. Our results originally describe mechanisms by which monocytes, a central cellular component recruited from peripheral blood to respiratory tract, succumb to control severe 2019 coronavirus disease (COVID-19).

**Author summary:** Since its emergence in China in late 2019, severe acute respiratory syndrome coronavirus 2 (SARS-CoV-2) has caused thousands of deaths worldwide. Currently, the number of individuals infected with SARS-CoV-2 and in need of antiviral, anti-inflammatory, anticoagulant and more invasive treatments has overwhelmed the health systems worldwide. In our study, we found that SARS-CoV-2 is capable of inducing inflammatory cell death in human monocytes, one of the main cell types responsible for anti-SARS-CoV-2 immune response. As a consequence of this intracellular inflammatory mechanism (inflammasome engagement), an exacerbated production of inflammatory mediators occurs. The infection also decreases the expression of HLA-DR in monocytes, a molecule related to the orchestration of the immune response in case of viral infections. We also demonstrated that the HIV-1 protease inhibitor, atazanavir (ATV), prevented the uncontrolled inflammatory response, cell death and reduction in HLA-DR expression in SARS-CoV-2-infected monocytes. Our study provides relevant information on the effects of SARS-CoV-2 infection on human monocytes, as well as on the effect of ATV in preventing these pathological effects on the host.

## Introduction

Severe acute respiratory coronavirus 2 (SARS-CoV-2), the etiological agent of the 2019 coronavirus disease (COVID-19), emerged in China, causing a major public health burden in decades. Patients with COVID-19 may develop an asymptomatic or mild disease or be affected by the life threatening acute respiratory distress syndrome (ARDS), which is characterized by elevated serum levels of proinflammatory mediators, the cytokine storm (1–3). In the respiratory tract of patients with severe COVID-19, monocytes/macrophages may be the main source of uncontrolled levels of the pro-inflammatory mediators TNF-α and IL-6 (4). Plasmatic levels of IL-6 have been associated with mortality, intensive care admission and hospitalization, representing a poor prognostic factor for COVID-19 (5). The uncontrolled inflammation promoted by SARS-CoV-2 in severe COVID-19 is not acute, it negatively associates with viral loads in nasopharyngeal swabs and represents an important event from 7 to 10 days after onset of illness (6,7). The SARS-CoV-2-induced cytokine storm associates with intense cell death (8,9).

There are various mechanisms involved in cell death, which are differently engaged from development to responses to infection (10). For certain diseases, such as COVID-19, in which the immunopathogenesis mechanisms associate with poor clinical outcomes, controlling the way cells collapse to infection is vital for the host (10). For example, cell death from necrosis, which can occur from necroptosis to pyroptosis, tends to exacerbate inflammation due to the rupture of the cellular plasma membrane. Pyroptosis, in particular, begins with the activation of the inflammasome, an intracellular structure that involves several intracellular molecules such as caspase-1(10,11), leads to leakage of the cytoplasmic content, favors inflammatory infiltrate (11) and amplifies of the inflammatory response (12). These mechanisms of cell death contrasts with apoptosis, a more controlled process that maintain the host’s homeostasis.

In severe COVID-19, the cytokine storm associates with high levels of tissue insult, judged by increased levels lactate dehydrogenase (LDH) and D-dimer in the plasma (6–8). Moreover, high LDH levels and leukopenia in severe COVID-19 points out that white cells loses the integrity of plasma membrane (6–8). Among these cells, monocytes should orchestrate the equilibrium between innate and adaptative immune responses, which may be presumably affected during cytokine storm. Indeed, severe immune dysregulation with low antigen presenting capacity is evidenced by reduced expression of HLA-DR in monocytes during COVID-19, which is strongly associated with severe respiratory failure (4).

The leukopenia of patients with severe COVID-19 seem to precede the cytokine storm (13–18). Moreover, in other virus-induced cytokine storm episodes in the respiratory tract, such as induced by influenza A virus, monocytes and macrophages are severely affected (13). Thus, we hypothesized that the monocyte cell death induced by SARS-COV-2 exacerbates the production of inflammatory cytokines, as well as impairs the immune balance in the hosts. In fact, we found that SARS-CoV-2 triggers the activation of the inflammasome, leading to pyroptosis in human monocytes, by experimental or natural infection. Pyroptosis was dependent on caspase-1 engagement, prior to IL-1ß production and dysregulation of cytokine release. Monocytes that survive after SARS-CoV-2 challenge downregulate HLA-DR, being less likely to properly orchestrate the immune response. Finally, we show that the reproposed antiviral drug atazanavir (ATV) could block this deleterious loop in favor of the host.

## Material and Methods

### Reagents

Atazanavir (ATV) and Ribavirin were received as donations from Instituto de Tecnologia de Fármacos (Farmanguinhos, Fiocruz). The antiviral Lopinavir/ritonavir (4:1 proportion) was pruchased from AbbVie (Ludwingshafen, Germany). ELISA assays were purchased from R&D Bioscience. Lipopolysacchadides - LPS, adenosine triphosphate (ATP), the specific inhibitor of caspase-1 (AC-YVAD-CMK), pan-caspase inhibitor (ZVAD-FMK), RIPK1 (Necrostatin-1 – Nec-1) and IL-1 receptor (IL-1RA) were all purchased from Sigma-Aldrich (St. Louis, MO, USA). All small molecule inhibitors were dissolved in 100 % dimethylsulfoxide (DMSO) and subsequently diluted at least 10^4^-fold in culture or reaction medium before each assay. The final DMSO concentrations showed no cytotoxicity. The materials for cell culture were purchased from Thermo Scientific Life Sciences (Grand Island, NY), unless otherwise mentioned.

### Cells and Virus

African green monkey kidney (Vero, subtype E6) cells were cultured in DMEM high glucose supplemented with 10 % fetal bovine serum (FBS; HyClone, Logan, Utah) and 100 U/mL penicillin, and 100 μg/mL streptomycin (P/S). Vero cells were incubated at 37°C in 5 % CO_2_ atmosphere.

Human primary monocytes were obtained through plastic adherence of peripheral blood mononuclear cells (PBMCs), which were obtained from buffy coat preparations of healthy donors by density gradient centrifugation (Ficoll-Paque, GE Healthcare). In brief, PBMCs (2.0 x 10^6^ cells) were plated onto 48-well plates (NalgeNunc) in RPMI-1640 without serum for 2 to 4 h; then, non-adherent cells were removed by washing and the remaining monocytes were maintained in DMEM with 5% human serum (HS; Millipore) and penicillin/streptomycin.

SARS-CoV-2 was isolated and expanded on Vero E6 cells from a nasopharyngeal swab of a confirmed case from Rio de Janeiro, Brazil. Experiments were performed after one passage in cell culture, when Vero E6 cells with DMEM plus 2% FBS in 150 cm^2^ flasks were incubated at 37 °C in 5 % CO_2_ atmosphere. Cytopathic effect was observed daily and peaked 4 to 5 days after infection. All procedures related to virus culture were handled at biosafety level 3 (BSL3) multiuser facility, according to WHO guidelines. Virus titers were determined as the tissue culture infectious dose at 50% (TCID50/mL). Virus stocks were kept in -80 °C ultralow freezers. The virus strain was sequenced to confirm the virus identity and its complete genome is publicly deposited (GenBank accession no. MT710714).

### Yield-reduction assay

Human primary monocytes were infected with multiplicity of infection (MOI) of 0.01 at density of 2-8 x 10^5^ cells/well in 48-well culture plates, depending on the total cell number from each donor. After 1 h at 37 °C, cells were washed, and various concentrations of compounds were added in DMEM with 2% FBS. After 48 h, the supernatants were harvested and virus replication was quantified by real time RT-PCR and infectious titers by TCID50/mL.

### Virus titration

Monolayers of Vero cells (2 x 10^4^ cell/well) in 96-well culture plates were infected with log-based dilutions of the supernatants containing SARS-CoV-2 for 1 h at 37°C. The cells were washed and fresh medium with 2% FBS was added. After 3 to 5 days, the cytopathic effects were scored in at least 10 replicates per dilution by independent readers, who were blind with respect to source of the supernatant. Reed and Muench scoring method was employed to determine TCID50/mL.

### Flow cytometer analysis

For flow cytometry analysis, monocytes were diluted in labelling buffer (10^6^ cells/mL). Then, 100 µL of cell samples were marked with 5 µL of AnnexinV and PI for 15 minutes for cell death evaluation. Around 10,000 gated events were acquired using FACSCalibur and the analysis was performed using the CellQuest software. Monocytes were gated through cell size (foward light scatter, FSC) and granularity (side light scatter, SSC) analysis.

Human monocytes were stained for caspase‐1 activity with FAM‐YVAD‐FMK (fluorescent‐labeled inhibitor of caspase‐1 [FLICA] and FAM-FLICA Caspase-3/7 activity or HLA-DR APC.H7 or IgG APC.H7. Caspase-1 and caspase-3/7 activity was determined via flow cytometry (FACSCalibur) by detecting FLICA fluorescence and expression of HLA-DR as mean fluorescence intensity (MFI) value for each sample. Acquisition of data was set to count a total of 10,000 events, and the FLOWJO software package was used to analyze the data.

### Microscopic analysis

Human primary monocytes were plated on glass coverslips at density of 2-8 x 10^5^ cells/well in 48-well plates. Infection was performed for 2 h at 37 °C and then fresh medium with 2% FBS was added. After 24 h, the cells were washed with Binding buffer and stained with PI (0.5 µg/mL) for 5 minutes. Next, the cells were fixed with 3.7% formaldehyde for 30 minutes at room temperature. The nuclei were stained with DAPI (1µg/mL) for 5 min and the coverslips were mounted using an antifade mounting medium (VECTASHIELD). Fluorescence was analyzed by fluorescence microscopy with an x100 objective lens (Olympus, Tokyo, Japan).

### Measurements Inflammatory Mediators and cell death marker

The levels of IL-1ß, TNF-α, IL-6, IL-8 and LDH were quantified in the culture supernatants from infected and uninfected monocytes using ELISA kits, according to the manufacturer’s intructions (R&D System).

Extracellular lactate dehydrogenase (LDH) was quantified using Doles^®^ kit according to manufacturer’s instructions. In brief, cell culture supernatants were centrifuged at 5,000 rpm for 1 minute, to remove cellular debris, and then 25 μL were placed into 96-well plates and incubated with 5 μL of ferric alum and 100 μL of LDH substrate for 3 minutes at 37 °C. Nicotinamide adenine dinucleotide (NAD, oxidized form) was added followed by the addition of a stabilizing solution. After 10 minutes, the reaction was read in a spectrophotometer at 492 nm.

### Western blot assay

Cellular extracts of 1×10^6^ cells were homogenized in the RIPA lysis buffer in the presence of proteinase inhibitor cocktail (Roche), and the protein levels were measured by BCA protein assay kit. A total of 20 μg of protein was loaded onto a 10% sodium dodecyl sulfate polyacrylamide gel (SDS-PAGE) for separation by electrophoresis and the protein bands were then transferred to a polyvinylidene difluoride membranes (ImmobilonP-SQ, Millipore). The membranes were blocked with 5 % albumin diluted in Tris-buffered saline containing 0.05 % of Tween 20 for 2 hours at room temperature and incubated with the specific primary antibodies (Cell Signaling Technology), to detect pro-caspase-1 and cleaved-caspase-1, after overnight incubation at 4 °C. After washing, membranes were incubated with secondary antibodies (IRDye® 800CW Goat-anti-Mouse and IRDye® 680LT Goat anti-Rabbit IgG Antibody, LI-COR, Lincoln) for 30 minutes at RT. The protein bands were visualized by digital fluorescence (Odyssey^®^ CLx Imaging System), and protein density was analyzed by the ImageJ software. All the data were normalized by β-actin expression quantification.

### Human subjects

We prospectively enrolled severe COVID19 RT-PCR-confirmed cases, as well as SARS-CoV-2-negative healthy controls. Blood samples were obtained from 12 patients with severe COVID-19 within 72 hours from intensive care unit (ICU) admission in two reference centers (Instituto Estadual do Cérebro Paulo Niemeyer and Hospital Copa Star, Rio de Janeiro, Brazil). Severe COVID-19 was defined as critically ill patients presenting viral pneumonia on computed tomography scan and in mechanical ventilation. All patients had SARSCoV-2 confirmed diagnostic through RT-PCR of nasal swab or tracheal aspirates. Peripheral vein blood was also collected from 8 SARS-CoV-2-negative healthy control participants as tested by RT-PCR on the day of blood sampling. The characteristics of severe (n = 12), and control (n = 8) participants are presented in Table Severe COVID-19 patients usually present older age and higher prevalence of comorbidities as obesity, cardiovascular diseases and diabetes as in previously reported patient cohorts. In the present study, the SARS-CoV-2-negative control group was designed to include subjects of older age and chronic non-communicable diseases, so it is matched with critically ill COVID-19 patients (Table 1). Patients with acute respiratory distress syndrome (ARDS) were managed with neuromuscular blockade and a protective ventilation strategy that included low tidal volume (6 mL/kg of predicted body weight) and limited driving pressure (less than 16 cmH2O) as well as optimal PEEP calculated based on the best lung compliance and PaO_2_/FiO_2_ ratio. In those with severe ARDS and PaO_2_/FiO_2_ ratio below 150 despite optimal ventilatory settings, prone position was initiated. Our management protocol included antithrombotic prophylaxis with enoxaparin 40 to 60 mg per day. Patients did not receive routine steroids, antivirals or other anti-inflammatory or anti-platelet drugs. The SARS-CoV-2-negative control participants were not under anti-inflammatory or anti-platelet drugs for at least two weeks. All clinical information were prospectively collected using a standardized form - ISARIC/WHO Clinical Characterization Protocol for Severe Emerging Infections (CCPBR).

**Table 1:** Characteristics of COVID-19 patients and control subjects.

| Characteristics<sup>1</sup> | Control<br>(N=8) | Covid-19<br>(N=9) |
| --- | --- | --- |
| Age, years | 52.6 (47 – 60) | 61 (33 – 79) |
| Sex, male | 8 (100 %) | 6 (66.7 %) |
| Respiratory support |  |  |
| Oxygen supplementation | 0 (0 %) | 3 (33.3 %) |
| Mechanical ventilation | 0 (0 %) | 6 (66.7 %) |
| SAPS 3 |  | 59.25 (31 – 75) |
| PaO<sub>2</sub>/FiO<sub>2</sub> ratio | - | 210.75 (87 – 509.5) |
| Vasopressor | - | 3 (33.3 %) |
| Time from symptom onset to<br>blood sample, days | - | 10 (6 – 18) |
| Status on Jun 30th |  |  |
| Dead | - | 5 (55.55 %) |
| Discharged | - | 1 (11.1 %) |
| Hospitalized | - | 3 (33.3 %) |
| **Comorbidities** |  |  |
| Obesity | 1 (11.1 %) | 1 (11.1 %) |
| Hypertension | 3 (33.3%) | 3 (33.3 %) |
| Diabetes | 0 (0%) | 0 (0 %) |
| Cancer | 0 (0%) | 0 (0 %) |
| Chronic heart disease<sup>2</sup> | 0 (0%) | 0 (0 %) |
| **Presenting symptoms** |  |  |
| Cough | 0 (0 %) | 3 (33,3 %) |
| Fever | 0 (0 %) | 3 (33,3 %) |
| Dyspnea | 0 (0 %) | 4 (44,4 %) |
| Headache | 0 (0 %) | 0 (0 %) |
| Anosmia | 0 (0 %) | 0 (0 %) |
| **Laboratory findings on**<br>**admission** |  |  |
| Lymphocyte count, cells/mm<sup>3</sup> | - | 1,355.8 (552 –2,564) |
| Platelet count, x 1000/mm<sup>3</sup> | - | 169.6 (92 – 278) |
| Leukocytes | - | 8,036 (8.03-18,670) |
| C Reactive Protein, mg/L | - | 14.78 (0.1 – 30.8) |

### Ethics statement

Experimental procedures involving human cells from healthy donors were performed with samples obtained after written informed consent and were approved by the Institutional Review Board (IRB) of the Oswaldo Cruz Foundation/Fiocruz (Rio de Janeiro, RJ, Brazil) under the number 397-07. The National Review Board approved the study protocol (CONEP 30650420.4.1001.0008), and informed consent was obtained from all participants or patients’ representatives.

### Statistical analysis

The assays were performed in blinded way. They were performed by one professional, codified and read by another fellow. All experiments were carried out at least three independent times, including a minimum of two technical replicates in each assay. The equations to fit the best curve were generated based on R^2^ values ≥ 0.9. Student’s T-test was used to access statistically significant P values <0.05.

## Results

### SARS-CoV-2 promotes pyroptosis in human monocytes

To characterize the mechanism by which SARS-CoV-2 triggers monocyte death, human primary monocytes were infected with SARS-CoV-2 or treated with LPS+ATP. Next, cell death was analyzed by quantifying the LDH activity in the culture supernatant, and by AnnexinV/propidium iodide (PI) cell labeling through for flow cytometry and fluorescence microscopy. As shown in Figure 1A, SARS-CoV-2 increased LDH levels similarly to the positive control, LPS+ATP (Figure 1A). Flow cytometry and fluorescence microscopy images demonstrated higher percentages of PI^+^ cells in the infected cultures, as well as a significant increase in mean fluorescence intensity (MFI). Both, LDH leak and PI labeling characterize cell membrane disruption in human monocytes after SARS-CoV-2 infection (Figure 1B-E). These data suggest that SARS-CoV-2 infection can induce cell death characterized by loss of plasma membrane integrity suggestive of pyroptosis cell death.

**Figure 1.**
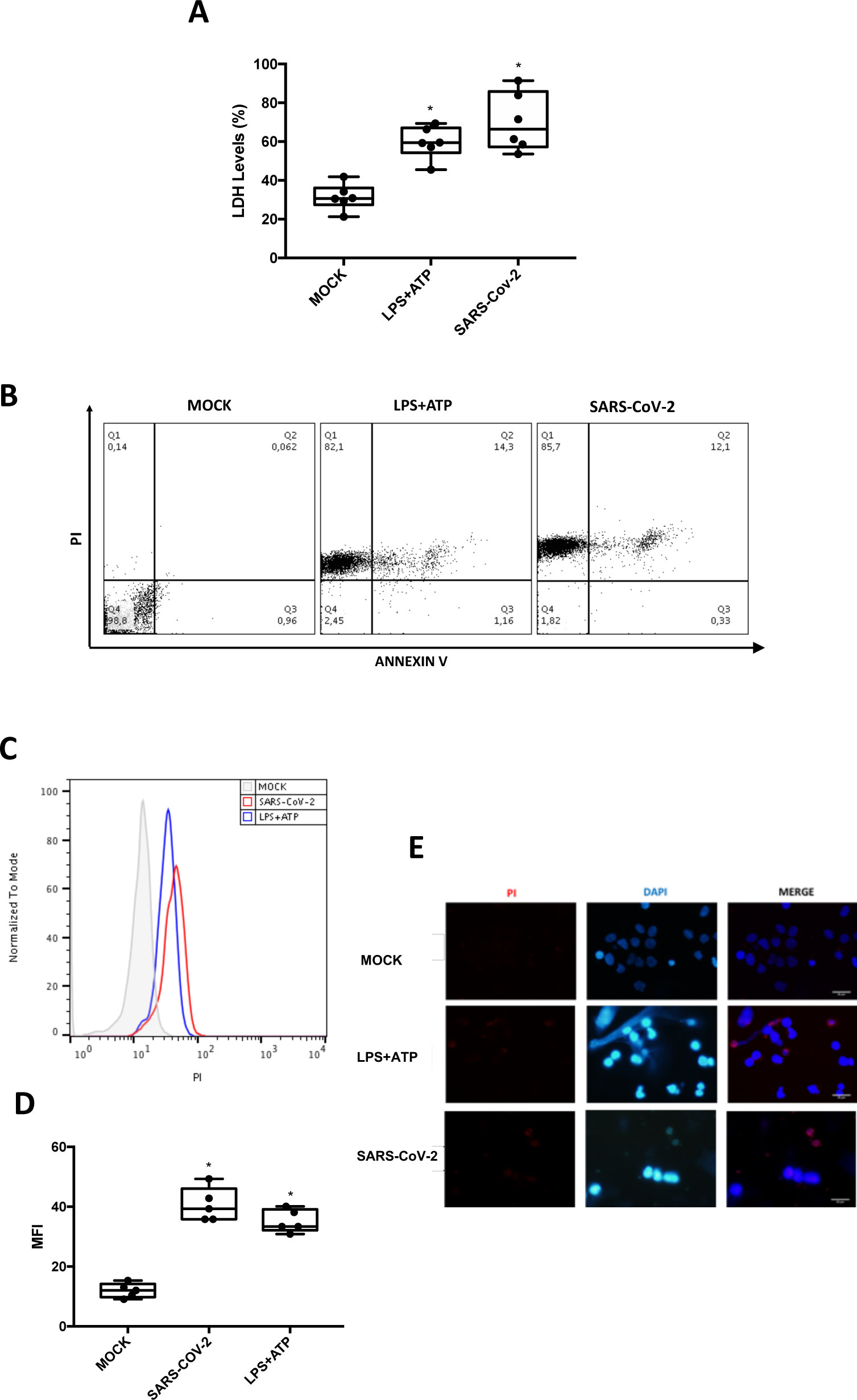
SARS-Cov-2 induces monocyte cell death through pyroptosis. Human monocytes were infected with SARS-CoV-2 (MOI 0.1) for 24 h. As a positive control, monocytes were stimulated with LPS (500ng/mL) for 23 h and after this time, incubated with ATP (2 mM) for 1 h. **(A)** Cell viability was assessed through the measurement of LDH release in the supernatant of monocytes. **(B, C and D)** Monocytes’ monolayers were stained with propidium iodide (PI) plus Annexin V and pyroptotic cell death were determine by flow cytometer analysis and **(E)** labeled with DAPI to visualize the nuclei fluorescence microscopy. Images and Graph data are representative of six independent experiments. Data are presented as the mean ± SEM ^*^ *P* < 0.05 *versus* control group (MOCK).

### SARS-CoV-2 induces pyroptosis in monocytes through inflammasome activation

To gain insights on the mechanisms of monocyte cell death in SARS-CoV-2 infection, we accessed caspase-1 activation - a key events that require inflammasome’s proteolytic activity. Cells stimulated with LPS+ATP were used as a positive control of inflammasome activation. We observed that SARS-CoV-2 induced pro-caspase-1 cleavage, similarly to positive control (Figure 2A and 2B). To confirm these results, cells were labeled with FAM-YVAD-FLICA (as an indicative of caspase-1 activation) and analyzed by flow cytometry. We found that SARS-CoV-2 infection induced the activation of caspase-1 in monocytes in the same magnitude to the positive control group (Figure 2C and 2D). Moreover, SARS-CoV-2-induced caspase-1 activation was a specific event, since we did not observe activation of the apoptotic caspases-3 and -7 in infected monocytes (Figure 2E and 2F). As caspase-1 activity is critical for the production of IL-1β (19,20), we measured the levels of this cytokine in our experiments. Our data show that SARS-CoV-2 infection was able to increase IL-1β production, this phenomenon being prevented with pretreatment with AC-YVAD-CMK, and not altered with a necroptosis inhibitor. (Figure 2G). Importantly, inhibition of IL-1R engagement reduced SARS-CoV-2-mediated caspase-1 activation and LDH release (Figure S1), suggesting that inflammasome-dependent IL-1β secretion amplify caspase-1 activation and pyroptosis in SARS-CoV-2 infection.

**Figure 2.**
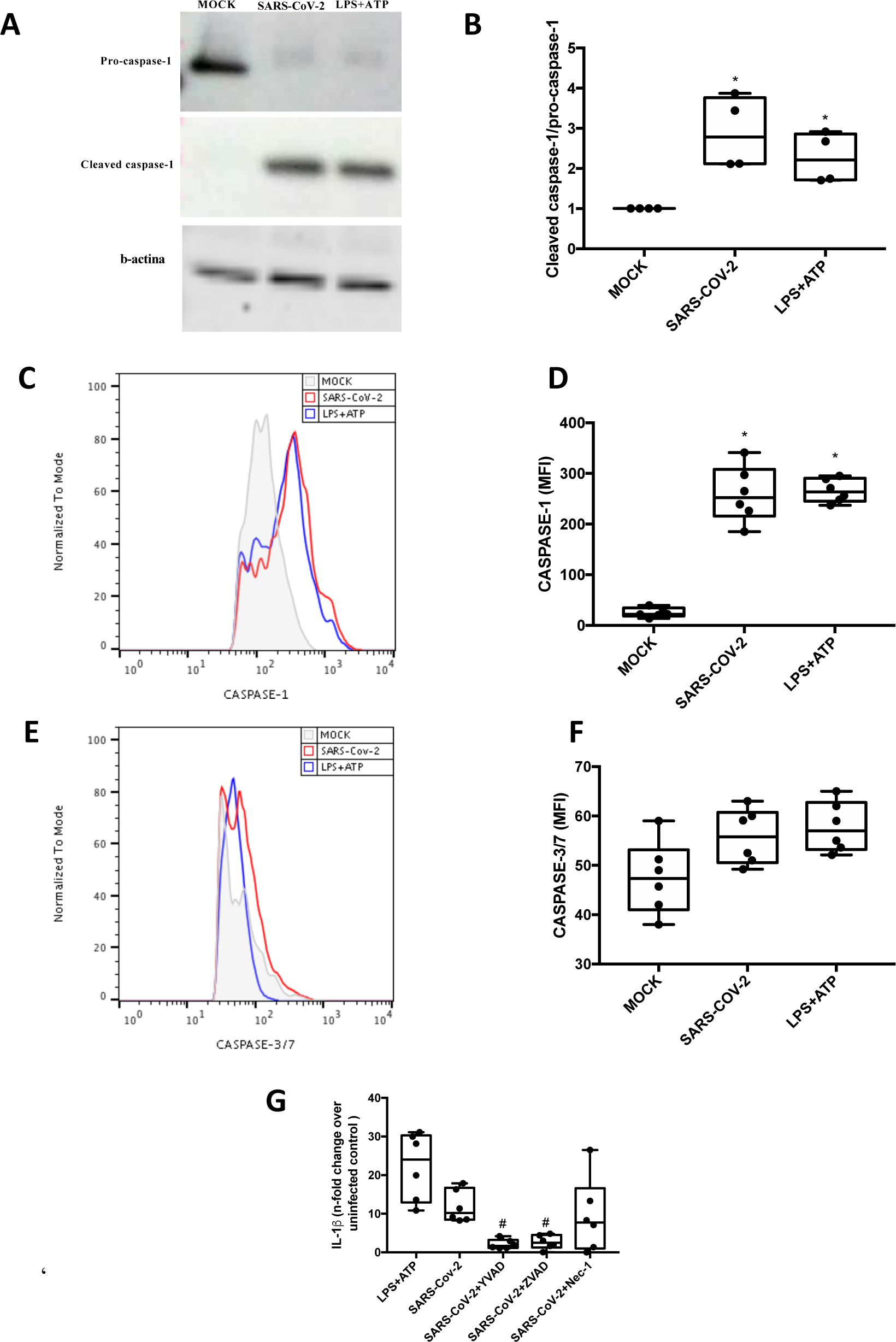
SARS-CoV-2 induce inflammasome activation in human monocytes. Human monocytes were pretreated with caspase-1 inhibitor (AC-YVAD-CMK – 1 µM), pan-caspase inhibitor (ZVAD-FMK - 10 µM) or necroptosis inhibitor (Nec-1 - 25 µM) for 1 h and infected with SARS-Cov-2 (MOI 0.1) for 24 h. Monocytes were stimulated with LPS (500ng/mL) for 23 h and after this time were stimulated with ATP (2 mM) for 1 h as a positive control group for the formation of inflammasomes and pyroptosis induction. **(A, B)** Monocytes’ monolayers were used for determination of the levels of pro-caspase-1 and cleaved caspase-1 by western blot analysis. **(C - F)** Monocytes were stained with FAM-YVAD-FLICA and FAM-FLICA to determine the caspase-1 and caspase-3/7 activity, respectively, by flow cytometry. **(G)** Cell culture supernatants were collected for the measurement of the levels of IL-1β. Western blot, histogram and graph data are representative of six independent experiments. Data are presented as the mean ± SEM ^#^ *P* < 0.05 *versus* infected and untreated group.

### Inflammasome activation amplify pro-inflammatory cytokines secretion in infected monocytes

To evaluate the effect of the activation of inflammasome and pyroptosis on the immune response, we quantified the production of key cytokines in the amplification of the inflammatory process observed clinically in a patient during the course of the infection with IL-6 and TNF-α. Remarkably, caspase-1 specific inhibition, but not pan-caspase or RIPK1 inhibitor, also led to lower levels of the pro-inflammatory cytokines IL-6 and TNF-α, highlighting the participation of the caspase-1-IL-1β axis in inflammatory amplification during SARS-CoV-2 infection. (Figure 3A and 3B). To confirm the effects in SARS-CoV-2 infection in the immune response, whether the observed effect is specifically related to inflammasome activation, we evaluated the production of IL-8, in which induction is independent of the inflammasomes pathway. SARS-CoV-2 infection induced a higher IL-8 production, which was not prevented by caspase-1 inhibition (Figure 3C). In addition, inhibition of IL-1R engagement reduced SARS-CoV-2-mediated was able to significantly decrease the production of IL-1 β, IL-6, TNF-α and did not alter the production of IL-8 (Figure S2).

**Figure 3.**
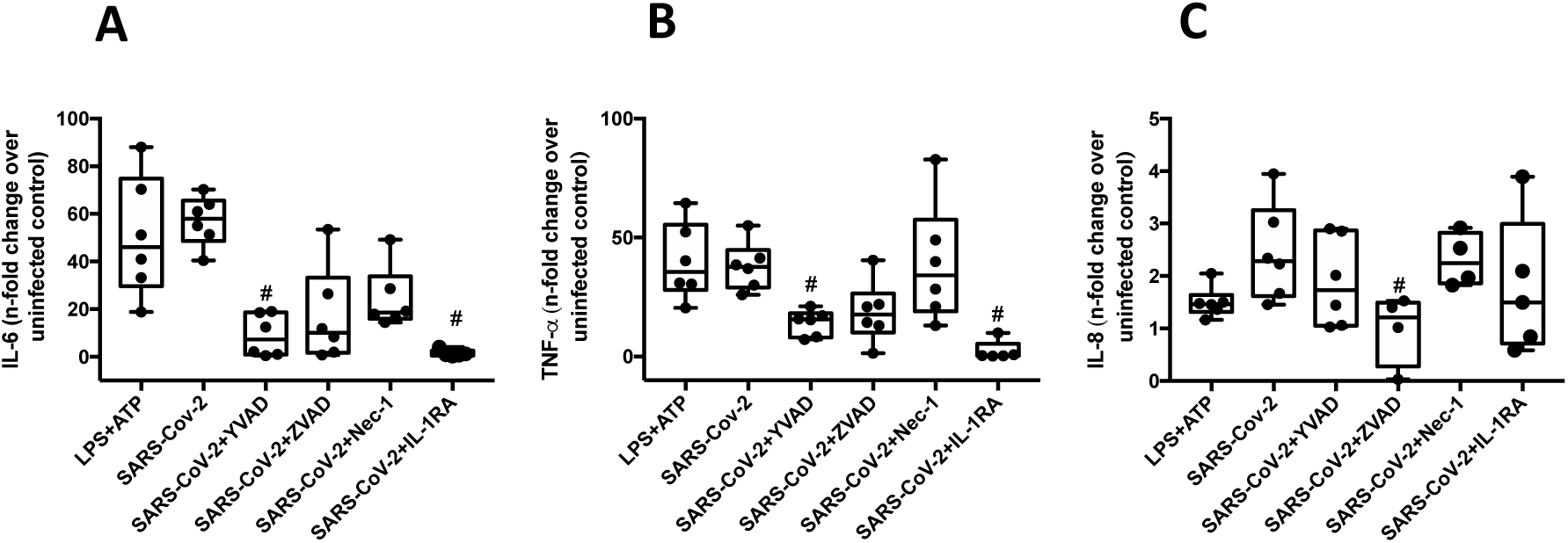
Inflammasome activation amplify pro-inflammatory cytokines secretion in infected monocytes. Human monocytes were pretreated with caspase-1 inhibitor (AC-YVAD-CMK – 1 µM), pan-caspase inhibitor (ZVAD-FMK - 10 µM), necroptosis inhibitor (Nec-1 - 25 µM) or with inhibitor of IL-1ß receptor (IL-1RA – 1 µM). Monocytes were stimulated with LPS (500ng/mL) for 23 h and after this time were stimulated with ATP (2 mM) for 1 h as a positive control group for the formation of inflammasomes and pyroptosis induction. Cell culture supernatants were collected for the measurement of the levels of **(A)** IL-6, **(B)** TNF-α and **(C)** IL-8. Graphs data are representative of six independent experiments. Data are presented as the mean ± SEM ^#^*P* < 0.05 *versus* infected and untreated group.

### Infection with SARS-CoV-2 decreases the expression of HLA class II in human monocytes independently of the activation of inflammasomes

To investigate the impact of inflammasome activation and cell death through pyroptosis on the orchestration of anti-SARS-CoV-2 immune response, cells were then marked with HLA-DR and analyzed by flow cytometry. We found that SARS-CoV-2 infection induced a decrease in HLA-DR expression in monocytes (Figure 4A and 4B). However, the inhibition of caspase-1 (Figure 4A and 4B) and IL-1R engagement (Figure S3) did not prevent the SARS-CoV-2-mediated decreasing of HLA expression. These results suggest that even in disassociation with the engagement of SARS-CoV-2-induced inflammasome activation, monocytes exposed to the new CoV will present limited ability to orchestrate the immune response.

**Figure 4.**
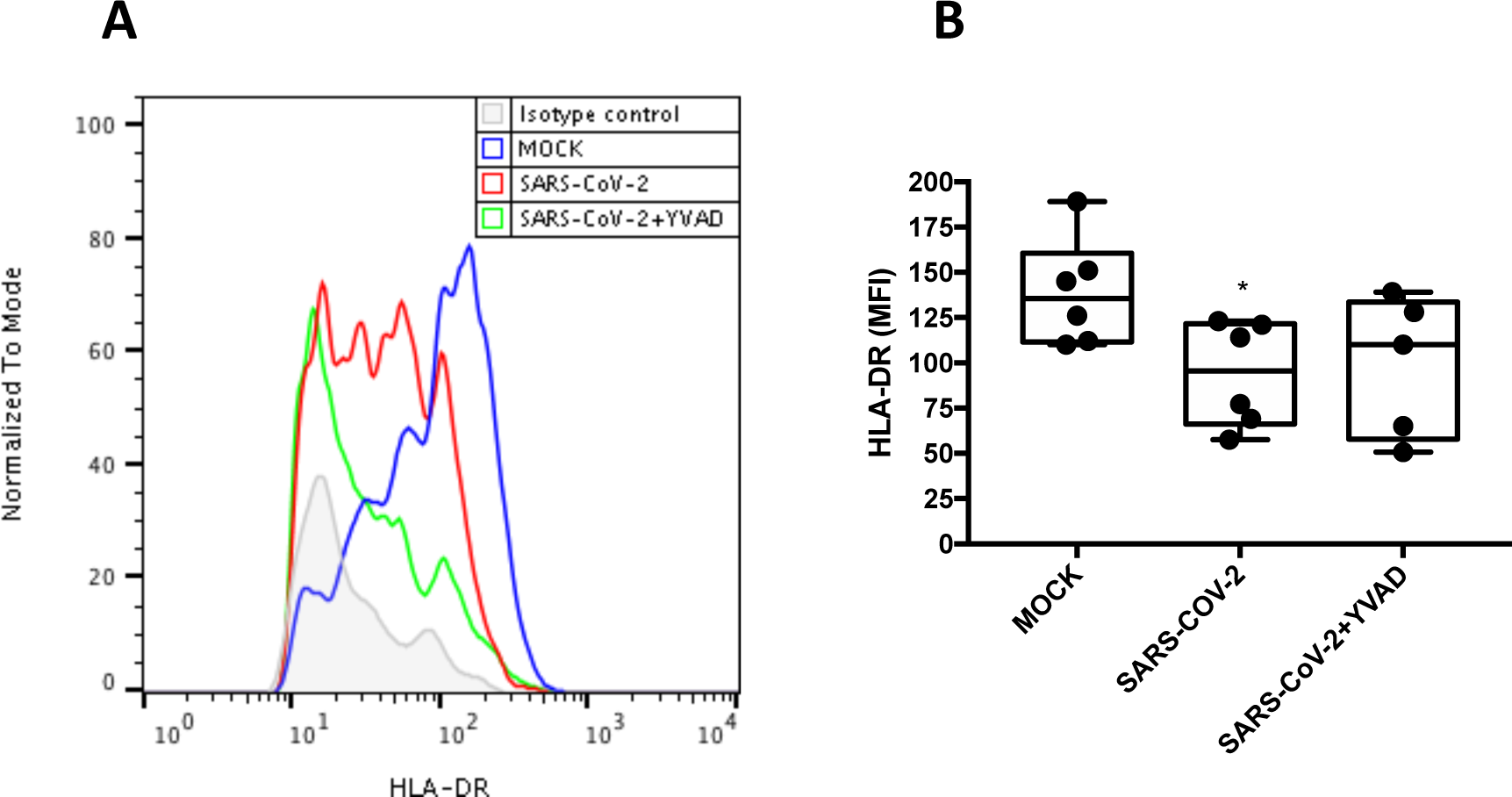
Infection with SARS-CoV-2 decreases the expression of HLA in human monocytes independently of the activation of inflammasomes. Human monocytes were pretreated with caspase-1 inhibitor (AC-YVAD-CMK – 1 µM) for 1 h and infected with SARS-Cov-2 for 24 h. **(A, B)** Monocytes were stained with HLA-DR APC.H7 or Isotype control APC.H7 to determine the HLA-DR expression by flow cytometry. Mean fluorescence intensity (MFI) value for each sample was represented in graphics. Histogram and graphs datas are representative of six independent experiments. Data are presented as the mean ± SEM ^*^ *P* < 0.05 *versus* uninfected group (MOCK).

### Atazanavir prevented inflammasome-dependent pyroptosis

Given our findings that pyroptosis and decreased HLA-DR expression are triggered during SARS-CoV-2 replication, we tested whether atazanavir (ATV), an HIV protease inhibitor recently shown to possess antiviral activity against the new CoV in monocytes (17), could prevent these events. Flow cytometry analysis of SARS-CoV-2-infected monocytes treated with ATV demonstrated a significant reduction in caspase-1 activity, observed by the decreased FAM-YVAD-FLICA labeling (Figure 5A and 5B). Other orally available repurposed anti-SARS-CoV-2 drugs, such as lopinavir (LPV) and ribavirin (RBV), did not affect SARS-CoV-2-induced pyroptosis (Figure 5C). Moreover, treatments with ATV did not alter the activity of caspase-1, -3 and -7 in monocytes exposed to LPS and ATP, used as a positive control of pyroptosis (Figure S4), indicating a specific antiviral activity of this drug. Consistently, treatment with ATV also reduced the levels of IL-1β, IL-6 and TNF-α in SARS-CoV-2-infected monocytes, when compared to the untreated cells (Figure 5D-F). ATV did not interfere with the production of IL-8, which is independent of inflammasome engagement (Figure 5G).

**Figure 5.**
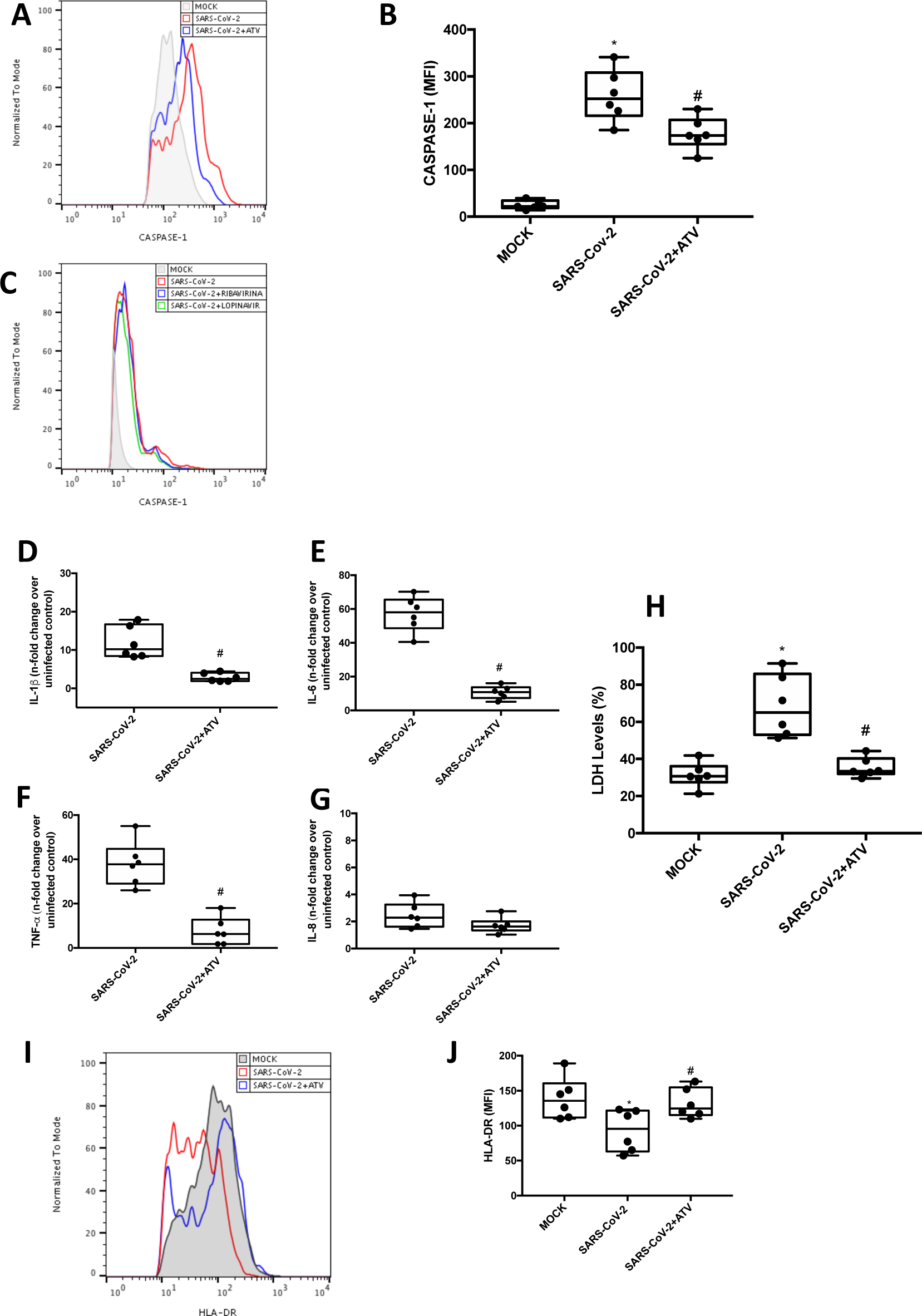
Atazanavir prevented inflammasome-dependent pyroptosis. Human monocytes were infected with SARS-Cov-2 and treated with atazanavir – ATV (10 µM), ribavirina (10 µM) or Lopinavir (10 µM) for 24 h. **(A, B and C)** Monocytes were stained with FAM-YVAD-FLICA or FAM-FLICA and HLA-DR APC.H7 or IgG APC.H7 to determine the caspase-1 and caspase-3/7 activity and HLA-DR expression, respectively and analyzed by flow cytometry. **(D – G)** Culture supernatants were collected and the levels of IL-1β, IL-6, TNF-α and IL-8 were determined by ELISA. **(H)** Assessment of cell viability through the measurement of LDH release in the supernatant of monocytes. **(I and J)** Monocytes were stained with HLA-DR APC.H7 to determine the HLA-DR expression by flow cytometry. Histogram and graphs datas are representative of six independent experiments. Data are presented as the mean ± SEM ^*^ *P* < 0.05 *versus* control group (MOCK); ^#^ *P* < 0.05 *versus* only infected group.

We then confirmed whether ATV can prevent SARS-CoV-2-induced pyroptosis. Lower levels of LDH were detected in the supernatants of SARS-CoV-2-infected monocytes treated with ATV, when compared to infected and untreated cells (Figure 5H). The treatment with ATV was also able to prevent the decrease of HLA-DR expression in infected monocytes (Figure 5I and J). LPV and RBV did not alter the HLA-DR expression in monocytes infected with SARS-Cov-2 (Figure S5). Since ATV inhibits the early proteolytic processing of viral antigens, an early event during SARS-CoV-2 replication, this drug represented the most upstream process pharmacologically inhibited in this investigation to prevent pyroptosis and HLA-DR knockdown.

### Inflammasome activation and pyroptosis are risk factors in critically ill COVID-19 patients

To clinically validate our findings, we evaluated if monocytes obtained from critically ill patients with COVID-19 would also display signals of inflammasome engagement and pyroptosis. We observed that monocytes from COVID-19 patients had increased caspase-1 activation (Figure 6A and 6B) and significantly higher PI+ staining, when compared to monocytes from healthy donors (HD) (Figure 6C and 6D). Corroborating with our *in vitro* data, we also detected higher levels of IL-1β in the plasma of critically ill patients (Figure 6E). Therefore, the *in vitro* results from the previous sections stand on the shoulders of the clinical relevance of monocytes in patients with severe COVID-19.

**Figure 6.**
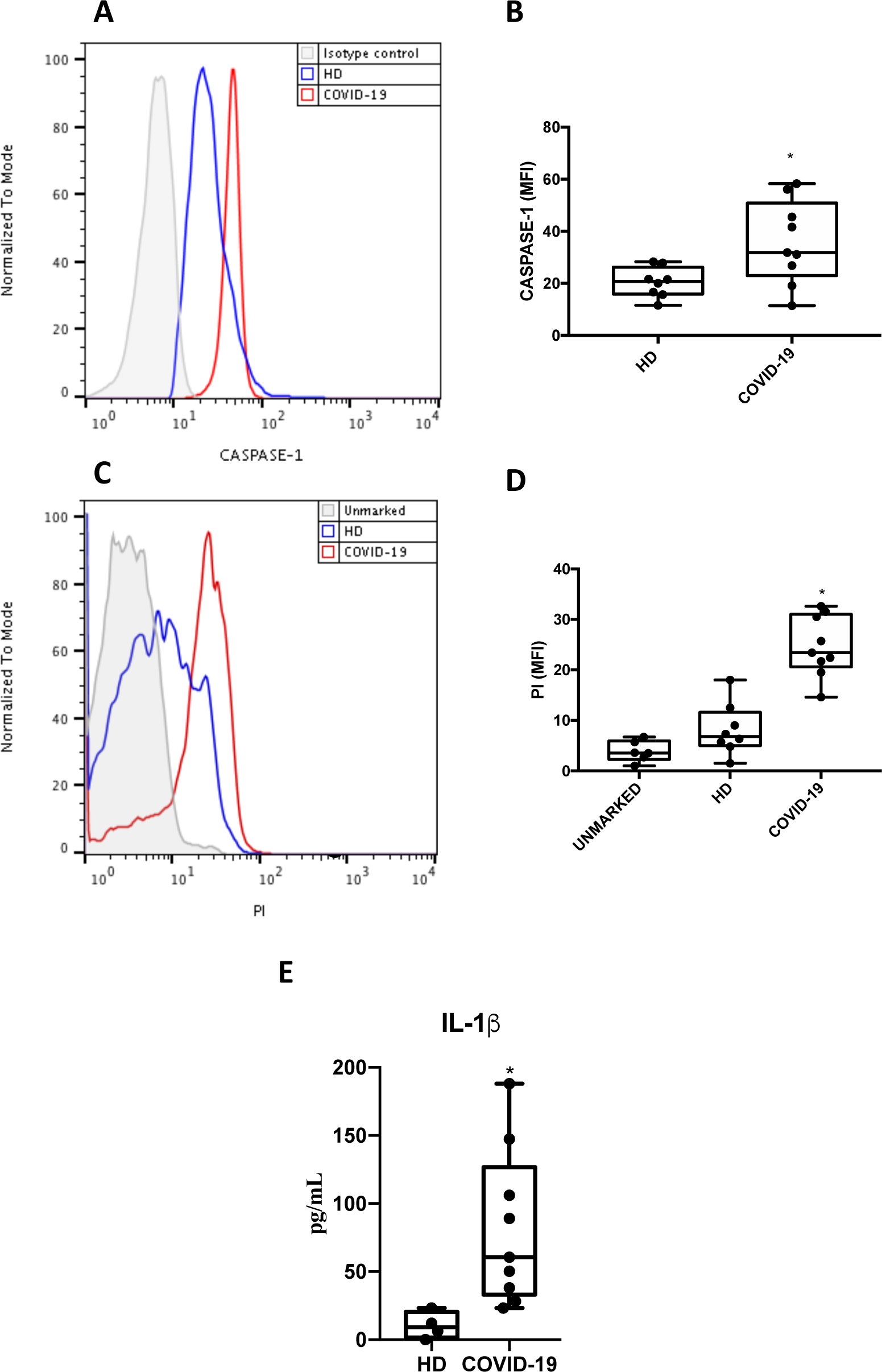
Inflammasome activation and pyroptosis are risk factors in critically ill COVID-19 patients. Monocytes isolated from blood samples of critically ill patients with COVID-19 and healthy donors. **(A and B)** monocytes were stained with FAM-YVAD-FLICA or **(C and D)** propidium iodide (PI) and analyzed by flow cytometry. **(E)** plasma was separated and the levels of IL-1β were determined by ELISA. Western blot, histogram and graphs datas are representative of 9 critically ill patients and 8 healthy donors. Data are presented as the mean ± SEM ^*^ *P* < 0.05 *versus* healthy donors (HD).

**Figure 7.**
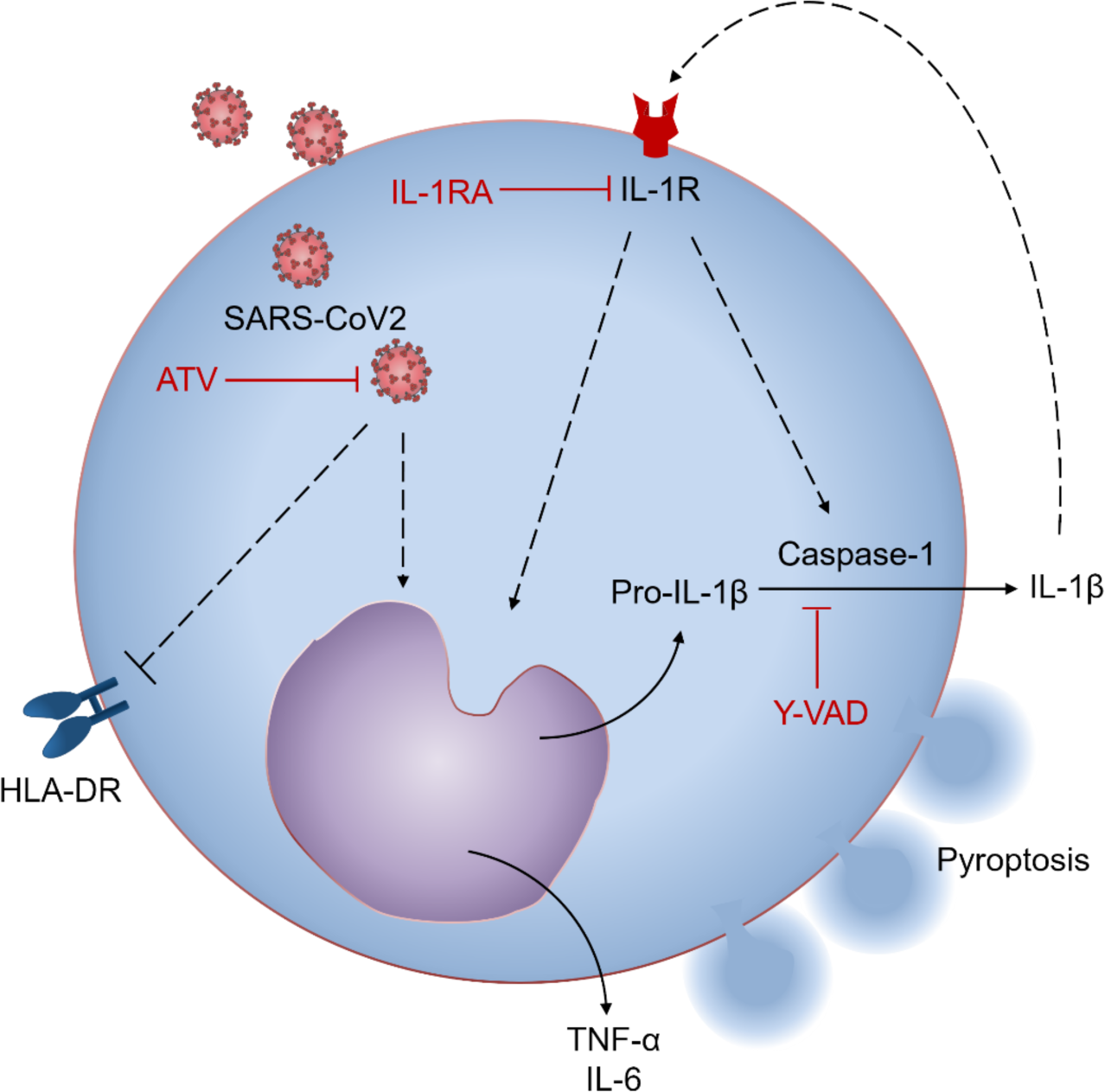
Representative scheme of the SARS-CoV-2 infection in activation of inflammasome and pyroptosis in monocyte with downregulation of HLA-DR expression and effects of ATV treatment in this phenomenon.

### SARS-CoV-2 infection induces inflammasome-dependent pyroptosis and downmodulation of HLA-DR monocytes, which can be prevented by atazanavir

A representative scheme to summarize the effect of SARS-CoV-2 infection upon in the increase of TNF-α and IL-6 levels, as part of an immunodysregulation promoted by pyroptosis. SARS-CoV-2-induced pyropotosis engages inflammasomes, to activate caspase-1 and IL-1ß. Release of IL-1ß amplify the activation of inflammasomes during the SARS-CoV-2 infection. In parallel, SARS-CoV-2 also induces the HLA-DR downregulation. ATV prevented SARS-CoV-2-induced deleterious effects on monocyte biology.

## Discussion

COVID-19 has caused in less than 8 months over 800,000 deaths worldwide (21) and represent the major public health crisis of the beginning of 21^st^ century, leading to an unpredictable impact in global economics (22,23). SARS-CoV-2 infection triggers an uncontrolled inflammatory response and marked leukopenia with consequent lung/respiratory dysfunction, which are the characteristics of the most severe manifestations of the COVID-19 (24,25). Similarly, to other respiratory viruses (26-29), SARS-CoV-2 induces a cytokine storm, characterized by an uncontrolled inflammatory response mediated by monocytes/macrophages, when they should orchestrate the antiviral immune response (30). In this work, we demonstrate that SARS-CoV-2 infection triggers inflammasome activation, increases IL-1ß secretion by monocytes, resulting in pyroptotic cell death, which could be a key event for SARS-CoV-2 pathogenesis in critically ill patients (31). This deleterious immune dysregulation loop triggered by SARS-CoV-2 may be impaired by ATV.

Our results demonstrate that SARS-CoV-2 leads to an intense cell death in human monocytes, observed by the increase in LDH release in infected cultures, as well as by the higher number of PI^+^ cells when compared to uninfected controls. Even though the pretreatment of infected monocytes with the pan-caspase inhibitor ZVAD-FMK prevented IL-1β secretion, it was not able to prevent the release of LDH. Indeed, others have demonstrated that the treatment with ZVAD-FMK induces necroptosis in diverse cell types (32-34). Notably, as pyroptosis is an inflammatory and programmed cell death mediated by inflammasome and inflammatory caspase-1 activation (10,11), we investigated the engagement and activation of these structures in human monocytes infected with SARS-CoV-2. Activation of caspase-1 and increased production of IL-1ß were observed in infected monocytes. This is correlated to the activation of inflammasomes, because the pretreatment of infected monocytes with YVAD prevented caspase-1 activation. These *in vitro* results are in accordance with the literature and with our finding described here that indicate the formation of inflammasomes in patients with severe COVID-19 (35). We also showed that the release of IL-1ß could be promoted by the activation of inflammasomes during the SARS-CoV-2 infection, because blockage of IL-1ß receptors reduced caspase-1 activation and cell death. These results corroborate with studies showing that the increase in IL-1ß production is associated with severe COVID-19 (36-38). Under our experimental conditions, the increase in IL-1ß precedes the unbalanced IL-6 release. These information should not be neglected when considering biopharmaceuticals to tackle cytokine storm in severe COVID-19.

Monocytes that survived from SARS-CoV-2 infection displayed decreased HLA-DR expression, which has also been described in monocytes isolated from COVID-19 patients (39,40). Our results suggest that this modulation of HLA expression is not directly associated with the activation of inflammasomes, and may be involved by other inflammatory pathways, since we and others have observed increased levels of IL-6 by myeloid cells impairing HLA-DR membrane expression (41).

To stablish the clinical relevance of SARS-CoV-2-induced pyroptosis in monocytes (41,41), we analyzed peripheral monocytes isolated from patients with severe COVID-19. We found that the cells from the patients displayed higher caspase-1 activation, when compared with monocytes isolated from HD. Recent clinical data reveal high levels of LDH and consistent leukopenia in critically ill COVID-19 patients (6,7,35,42-44). Our data also demonstrate intense monocyte death in COVID-19 patients, as detected through flow cytometry analyzes. Altogether, these data suggest that the severity of COVID-19 may be associated with inflammasome activation in monocytes that results in large amounts of IL-1ß and generates an excessive inflammatory response, further characterized by high levels of IL-6 and TNF-α. Consistently, treatment with IL-1RA has been associated with clinical and inflammatory improvements in critically ill COVID-19 patients (45). These results are in line with clinical case reports that demonstrate that monocytes/macrophages are key cells in the deleterious pro-inflammatory events that characterize the most serious cases of COVID-19 (46-49).

To the best of our knowledge, effective therapies for COVID-19 should ideally combine antiviral/anti-inflammatory drugs to reduce viral load and to mitigate the cytokine storm. In the present study, we showed that ATV, an antiretroviral approved in 2003 for HIV-1 treatment and previously described by us as having anti-SARS-CoV-2 effects in different cell types (including monocytes)(50), to block new CoV-induced pyroptosis and HLA-DR knockdown in monocytes. In addition, ATV inhibited the release of LDH and the production of IL-1ß, f IL-6 and TNF-α from SARS-CoV-2-infected monocytes, which are key players in the cytokine storm associated with COVID-19 (51,52).

In this work, we originally describe that infection with SARS-CoV-2 can induce pyroptotic cell death by inflammasome activation, which may be related to the intense leukopenia and exacerbated inflammation seen in severe cases of the COVID-19. Since there is no specific therapy for this disease, our results point out that ATV has a promising therapeutic potential against SARS-CoV-2-induced cell death.

## Data Availability

The data supporting the results of this study are available as supplementary files on the website mendeley.com

## Acknowledgments and funding

We thank the Hemotherapy Service from Hospital Clementino Fraga Filho (Federal University of Rio de Janeiro, Brazil) for providing buffy-coats. This work was supported by grants from Fundação de Amparo a Pesquisa do Estado do Rio de Janeiro (FAPERJ), Inova Fiocruz, Conselho Nacional de Desenvolvimento Científico e Tecnológico (CNPq) and Coordenação de Aperfeiçoamento de Pessoal de Nível Superior (CAPES) and Mercosur Fund for Structural Convergence (FOCEM, Mercosur, grant number 03/11) granted for Thiago Moreno L. Souza, Patrícia T. Bozza, Fernando A. Bozza and Dumith Chequer Bou-Habib. Funding was also provided by CNPq, CAPES and FAPERJ through the National Institutes of Science and Technology Program (INCT) to Carlos Morel (INCT-IDPN) and to Wilson Savino (INCT-NIM). Thanks are due to Oswaldo Cruz Foundation/Fiocruz under the auspicious of Inova program. The funding sponsors had no role in the design of the study; in the collection, analyses, or interpretation of data; in the writing of the manuscript, and in the decision to publish the results.

## FIGURE LEGENDS

**S1 Fig.**
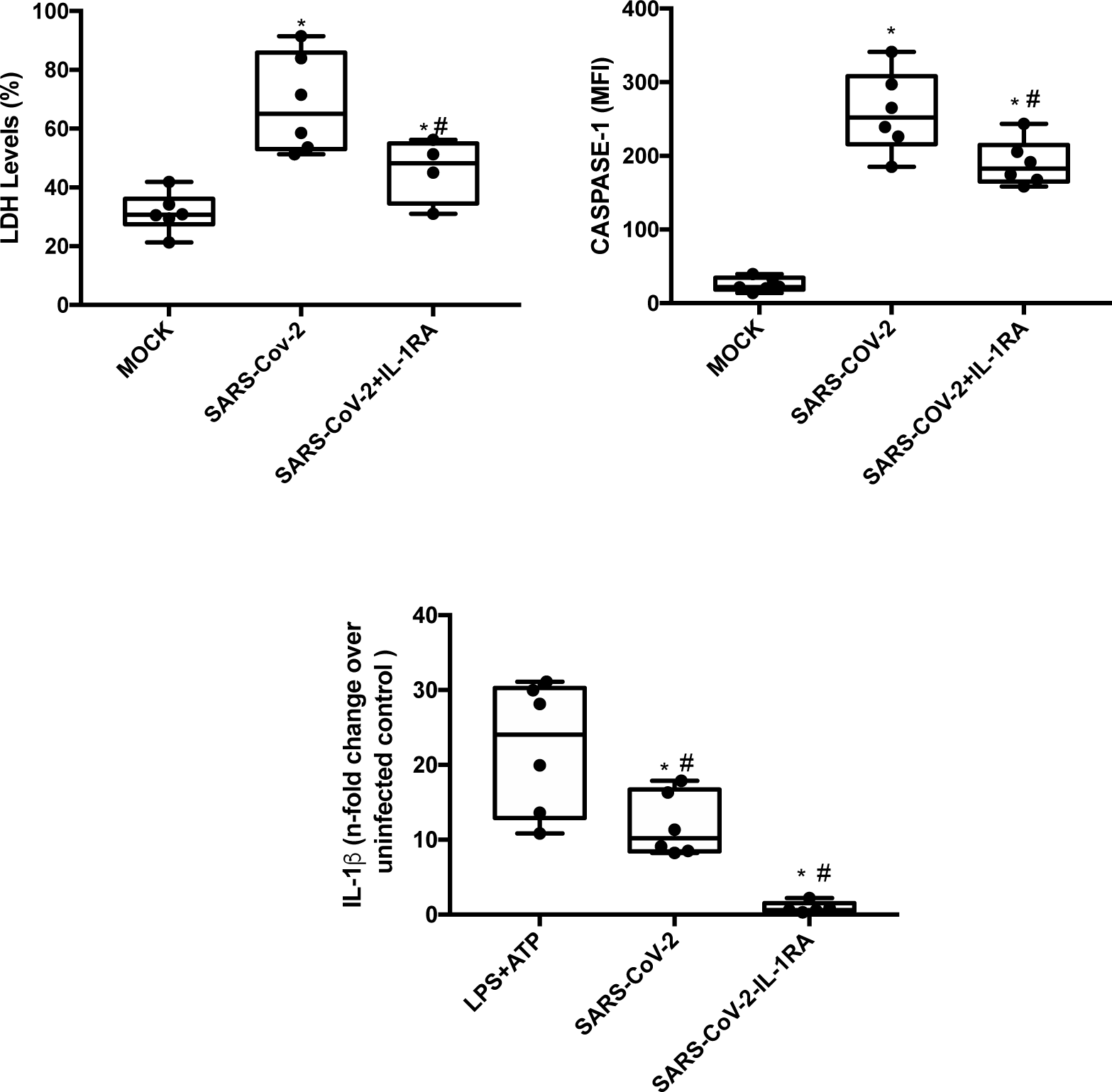
SARS-CoV-2 induce inflammasome activation and pyroptosis in human monocytes by IL-1β amplification. Human monocytes were pretreated with IL-1β receptor (IL-1RA – 1 µM) for 1 h and infected with SARS-Cov-2 (MOI 0.1) for 24 h. Cell viability was assessed through the measurement of LDH release in the supernatant and monocytes were stained with FAM-YVAD-FLICA to determine the caspase-1 and caspase-3/7 activity, respectively, by flow cytometry. Cell culture supernatants also were collected for the measurement of the levels of IL-1β. Data are presented as the mean ± SEM ^#^ *P* < 0.05 *versus* infected and untreated group; ^*^ *P* < 0.05 *versus* control group (MOCK). Graphs are representative of four independent experiments.

**S2 Fig.**
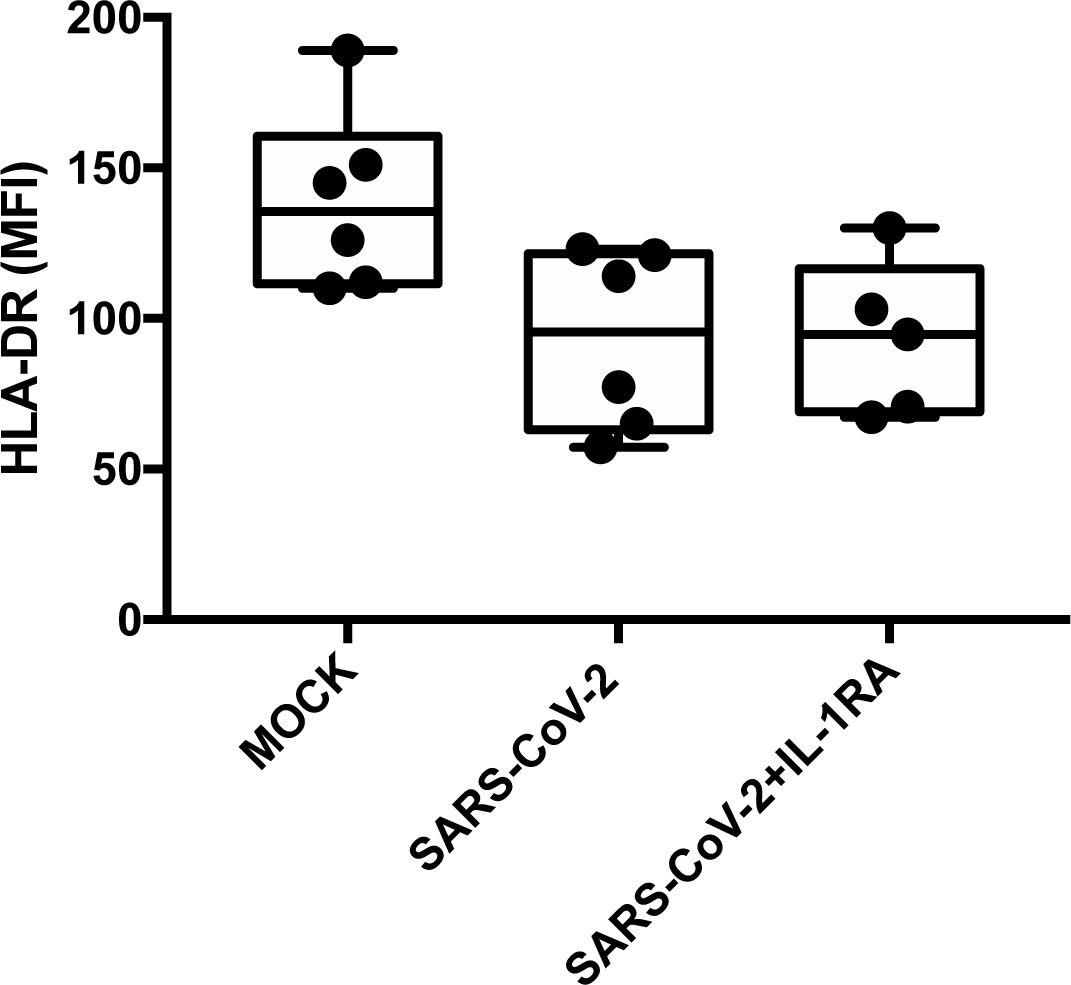
Infection with SARS-CoV-2 decreases the expression of HLA in human monocytes independently of the IL-1ß activation. Human monocytes were pretreated with inhibitor of IL-1ß receptor (IL-1RA – 1 µM) for 1 h and infected with SARS-Cov-2 for 24 h. Monocytes were stained with HLA-DR APC.H7 or IgG APC.H7 to determine the HLA-DR expression by flow cytometry. Mean fluorescence intensity (MFI) value for each sample was represented in graphics. Graphs datas are representative of four independent experiments. Data are presented as the mean ± SEM ^#^ *P* < 0.05 *versus* uninfected group (MOCK).

**S3 Fig.**
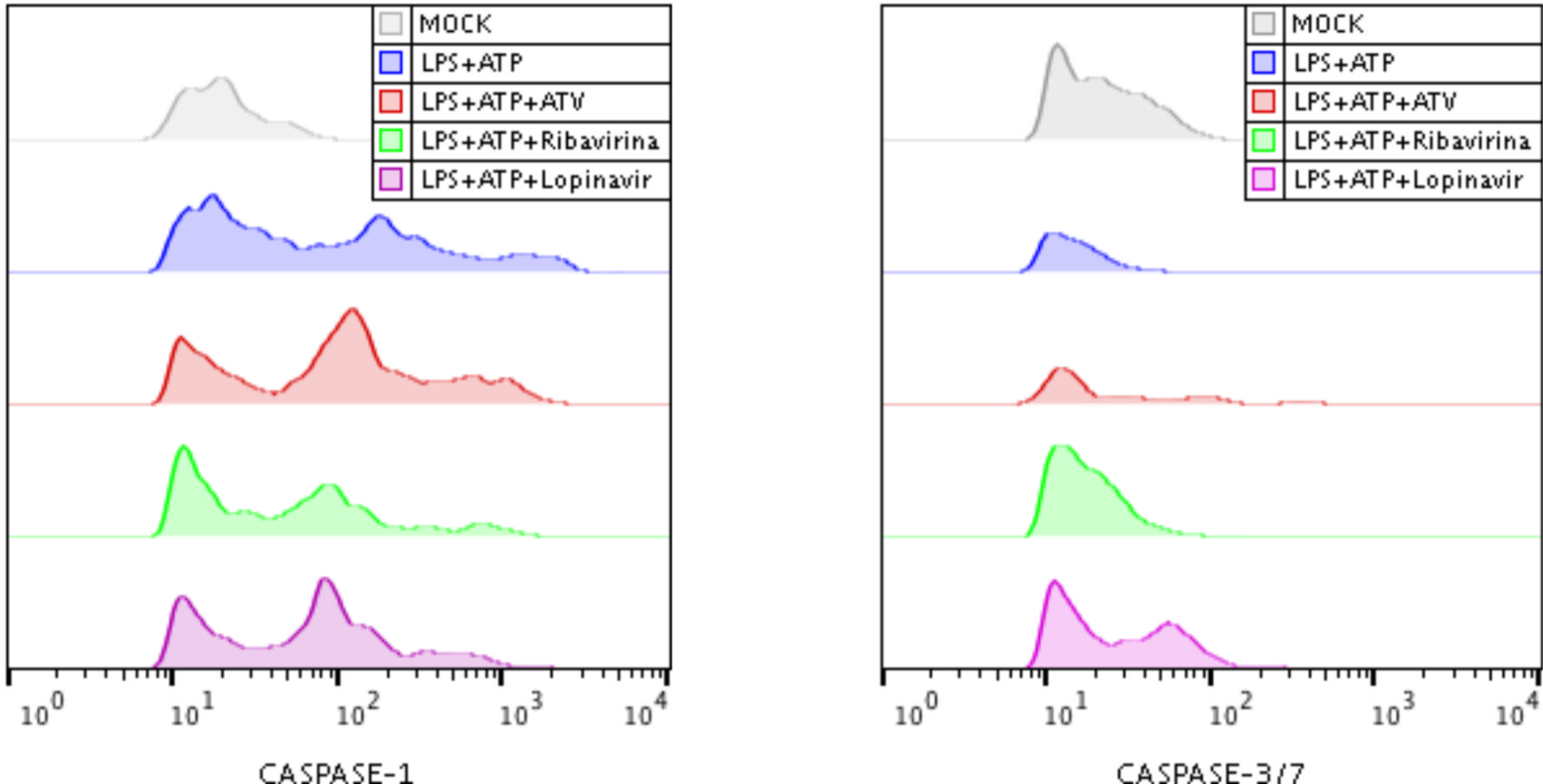
Evaluation of anti-inflammatory activity of ATV, LPV and ribavirin in LPS-induced monocytes. Human monocytes were stimulated with LPS (500ng/mL) for 23 h and after this time were stimulated with ATP (2 mM) for 1 h as a positive control group for the formation of inflammasomes and pyroptosis induction. Simultaneously, cells were treated with atazanavir – ATV (10 µM), LPV (10 µM) or ribavirin (10 µM) for 24 h. Monocytes were stained with FAM-YVAD-FLICA (blue) or FAM-FLICA (green) to determine the caspase-1 and caspase-3/7 activity and HLA-DR expression, respectively, and analyzed by flow cytometry. Histograms are representative of six independent experiments.

**S4 Fig.**
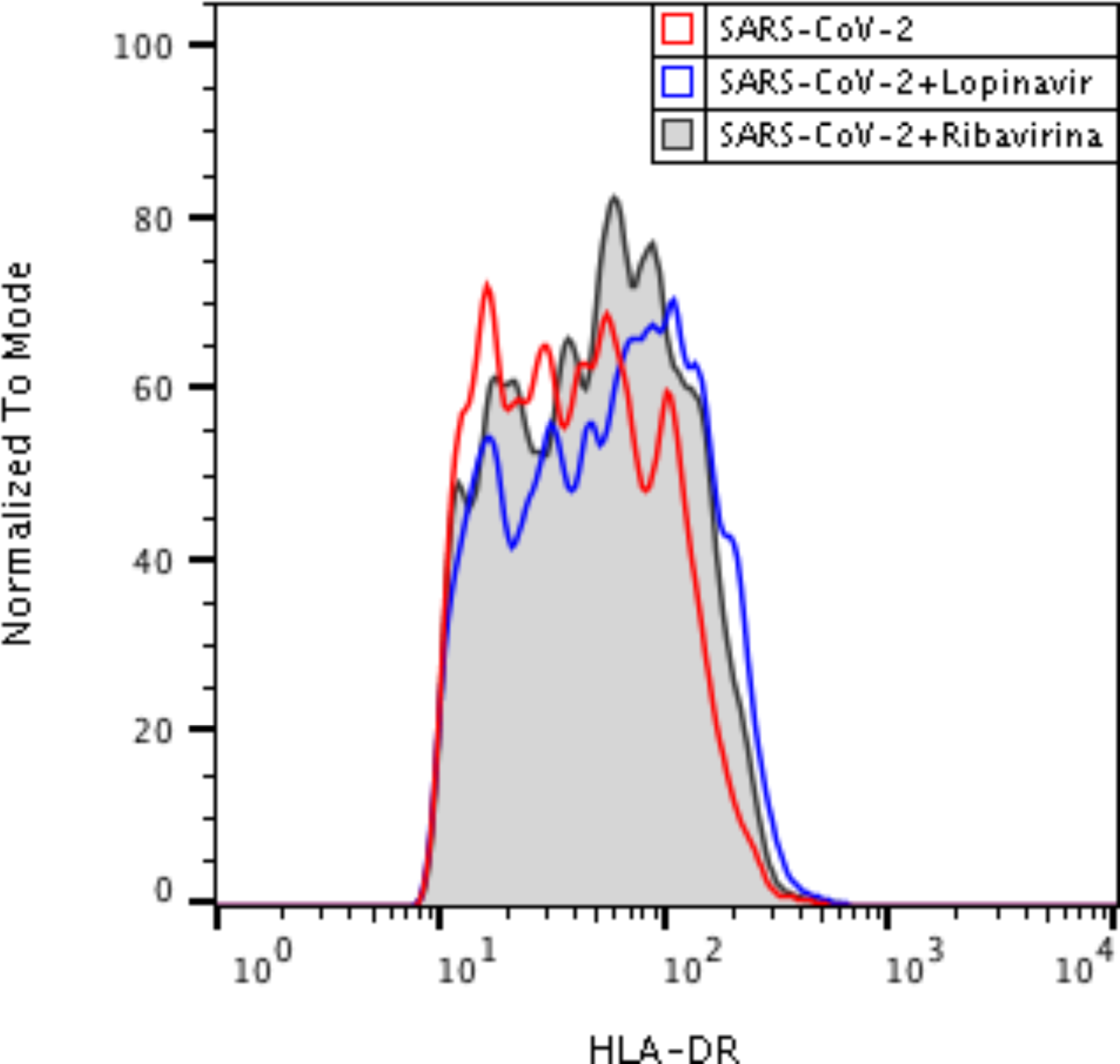
Effects of LPV and ribavirin in preventing the SARS-CoV-2-related reduced expression of HLA-DR in monocytes. Human monocytes were infected with SARS-Cov-2 and treated with LPV (10 µM) or ribavirina (10 µM) for 24 h. Monocytes were stained with HLA-DR APC.H7 or IgG APC.H7 to determine the HLA-DR expression by flow cytometry. Histogram data are representative of six independent experiments.

